# Age Differences in the Reproducibility of Seasonal Peak Timing for Alcohol-Associated Injury: A Seven-Year Cosinor and Jackknife Analysis of U.S. Emergency Department Surveillance Data

**DOI:** 10.64898/2026.08.27.26361527

**Authors:** Devpal Ghuman, Tarun Achar, Dhruv Gambhirrao

**Affiliations:** Texas Academy of Mathematics and Science, University of North Texas, Denton, TX, United States of America

**Keywords:** Alcohol-related injury, Emergency department, Seasonality, Age factors, Sex differences, Injury surveillance, Cosinor analysis, Older adults

## Abstract

**Background:** Alcohol-associated injury is a leading cause of emergency department (ED) utilization in the United States and a clinically important driver of preventable morbidity across the adult lifespan. Prior surveillance research has characterized how the rate and severity of alcohol-associated injury vary by patient age, but whether the seasonal timing of injury risk is equally predictable across age groups (a question directly relevant to the timing of clinical screening intensification and public health intervention) has not been formally tested.

**Methods:** We conducted a retrospective surveillance analysis of 45,876 alcohol-associated ED visits among adults aged 18 years and older, identified from the National Electronic Injury Surveillance System (NEISS), 2019-2025 (weighted national estimate: 2,092,319 visits), using the structured Alcohol_Involved indicator introduced into NEISS case abstraction in 2019. Patients were stratified by sex and five age groups (18-24, 25-34, 35-49, 50-64, and ≥65 years). Single-harmonic cosinor (Poisson) regression was used to estimate the seasonal peak day of injury risk (acrophase) for each stratum. To assess reliability, we performed leave-one-year-out jackknife resampling (seven iterations per group), case-resampling bootstrap confidence intervals (1,000 iterations), and likelihood-ratio tests of seasonal-phase interactions.

**Results:** Peak injury timing differed significantly across age groups (X^2 [8] = 2356.2, p < .0001). Adults aged 25-64 years showed a highly reproducible early-to-mid-July peak, with jackknife estimates shifting ≤14 days when any single study year was excluded. Adults aged ≥65 years showed significant seasonal variation annually (all p < .0001, amplitude comparable to younger groups) but a pooled peak estimate that shifted by up to 100 days across jackknife iterations. Sex-stratified analyses revealed that this instability was driven entirely by females aged ≥65 years (jackknife range: 332 days, peak consistently in late October through early January) rather than males aged ≥65 (jackknife range: 31 days, peak consistently in early August). Hospital admission rates increased monotonically with age from 9.0% (18-24 years) to 31.8% (≥65 years).

**Conclusions:** Alcohol-associated injury follows a reproducible, calendar-stable summer seasonal pattern in adults aged 25-64 years. Among adults ≥65 years, the previously reported temporal instability is concentrated in the female subgroup, whose seasonal injury risk does not converge on a fixed calendar window. These findings suggest that fixed-calendar prevention and screening strategies are well suited to working-age adults and older men, but older women may require a year-round, individually tailored approach.

## INTRODUCTION

Alcohol use is a well known risk factor for accidental and intentional injury seen in the emergency department (ED). This includes falls, traffic crashes, assaults, and burns (Cherpitel, 2007). Injuries related to alcohol use have a significant impact on clinical practice and public health as they can increase ED overcrowding, frequency of hospitalizations, and costs to the trauma system (Rehm et al., 2009). In the United States alone, approximately 95,000 deaths per year are attributed to excessive alcohol use, making it the third leading preventable cause of death (Centers for Disease Control and Prevention, 2022).

Prior studies identified how alcohol injuries differ by patient age. CDC studies of nationally collected ED data have documented growing rates of alcohol-associated falls among older patients over the past decade (Centers for Disease Control and Prevention, 2022). In this group specifically, age-related physical changes, polypharmacy, and comorbidities contribute to increased risk and severity of alcohol-related trauma. Older adults are particularly vulnerable because alcohol metabolism slows with age, lower doses produce higher blood alcohol concentrations, and many commonly prescribed medications interact adversely with alcohol to increase fall risk (Saitz, 2005). Sex differences in alcohol metabolism are also well established: women develop higher blood alcohol concentrations than men after equivalent alcohol doses due to lower total body water and lower gastric alcohol dehydrogenase activity (Nolen-Hoeksema, 2004). These physiological differences may manifest in distinct seasonal patterns of alcohol-related injury across both age and sex strata.

The separate field of chronobiology has revealed circadian and seasonal patterns in alcohol consumption behavior at both the individual and population level (Manzardo, Poje, & Penick, 2021; Merikangas et al., 1990). Population-level studies have documented seasonal peaks in alcohol-related motor vehicle crashes, emergency department presentations, and hospitalizations, with most research identifying summer as a period of elevated risk (Smith & Crum, 1999). However, whether the timing of alcohol-related injury risk in older adults and in women follows the same seasonal pattern as younger adults and men has not been tested. This matters because the physiology of alcohol metabolism, the social contexts of drinking, and the circumstances leading to injury all differ meaningfully by age and sex.

To our knowledge, no previous research has specifically tested whether the seasonal timing, as opposed to the rate, of alcohol-related injuries is equally predictable across age and sex groups using nationally representative U.S. ED data. This is both an important clinical and operational question. A seasonal peak in injury occurrence can only be used for clinical and public health applications if it recurs every year around the same time. We refer to this distinction as reproducible versus non-stationary seasonality, and we test it using cosinor regression applied to seven years (2019–2025) of U.S. ED surveillance data, with leave-one-year-out jackknife resampling as a novel robustness test.

## METHODS

### Study Design and Data Source

We conducted a retrospective, repeated cross-sectional surveillance analysis using the National Electronic Injury Surveillance System (NEISS), operated by the U.S. Consumer Product Safety Commission (CPSC) in partnership with the Centers for Disease Control and Prevention. The NEISS data are probability samples of injury visits from a representative panel of approximately 100 hospital EDs throughout the country, with case-based statistical weights provided to enable national estimates of ED visit counts. Our analysis used the publicly available yearly NEISS case-based microdata for treatment years 2019 through 2025 (7 years; N = 2,442,005 total injury cases).

### Study Population

All eligible cases were ED visits among patients aged 18 years and older where there was evidence of alcohol involvement, as determined by the structured Alcohol_Involved indicator field included in NEISS data abstraction for all treatment dates starting January 1, 2019. This field is coded by trained NEISS abstractors based on documented clinical evidence of alcohol involvement at the time of injury (e.g., blood alcohol level, clinician documentation of intoxication, or patient report of alcohol use in the medical chart). Using the structured field without supplementary free-text narrative classification avoids the misclassification associated with text-mining approaches required in NEISS years before 2019. During the study period, 45,876 of 1,441,585 adult injury records (weighted: 2,092,319 of an estimated 59.4 million adult ED visits) had documented alcohol involvement.

Subjects were stratified by five age groups (18–24, 25–34, 35–49, 50–64, and ≥65 years) and by sex (male, female), consistent with standard epidemiological life-stage categories and NEISS coding conventions. Cases with unknown sex (n = 16, weighted = 736) were excluded from sex-stratified analyses. Hospital admission was defined as NEISS disposition code 4 (treated and admitted/hospitalized) and was calculated by age group as a clinical severity indicator.

### Statistical Analysis

For each age group and sex stratum, weighted daily case counts were aggregated across all seven study years by day of year (1–366, retaining February 29 in leap years). We fit a single-harmonic cosinor model via Poisson regression:

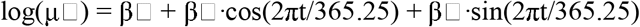

where t indexes day of year and μ□ is the expected daily weighted case count. The fitted coefficients yield the amplitude (A = √[β□^2^ + β□^2^]) and the acrophase (peak day of year). The seasonal component was tested via likelihood ratio test against an intercept-only null model for each stratum and each individual study year.

Estimate reliability was evaluated using three approaches. First, 95% confidence intervals for the pooled acrophase were estimated by case-resampling bootstrap (1,000 resamples), using circular statistics appropriate for cyclical day-of-year data. Second, pairwise bootstrap comparisons tested whether peak timing of each group differed significantly from the 25–34-year reference group (selected post hoc as the most stable group). Third, leave-one-year-out jackknife resampling (seven iterations per stratum) yielded jackknife peak-day estimates; the jackknife range (maximum − minimum) is reported as a year-over-year reproducibility measure, conceptually distinct from sampling-based confidence intervals.

A sensitivity analysis excluded 2020 (disrupted by COVID-19) and compared results across early (2019–2021) and late (2022–2025) sub-periods. All analyses were performed in Python 3.12 using statsmodels and scipy. As this study used publicly available, de-identified secondary surveillance data, institutional review board approval was not required.

## RESULTS

### Sample Characteristics

The analytic sample comprised 45,876 alcohol-associated ED visits among adults aged 18 and older (weighted national estimate: 2,092,319 visits, 2019–2025). By sex, 31,077 cases (67.7%; weighted: 1,404,547) were male and 14,783 (32.2%; weighted: 687,036) were female. Weighted visits by age group were: 18–24 years, 221,930; 25–34 years, 338,687; 35–49 years, 460,978; 50–64 years, 630,100; and ≥65 years, 440,623. All five age groups showed statistically significant seasonal variation in every individual study year (likelihood ratio test, all p < .0001). Hospital admission rates increased monotonically with age, from 9.0% (18–24 years) to 13.0%, 19.4%, 25.3%, and 31.8% (≥65 years), which is a more than 3-fold gradient in injury severity (Table 1).

**Table 1.**
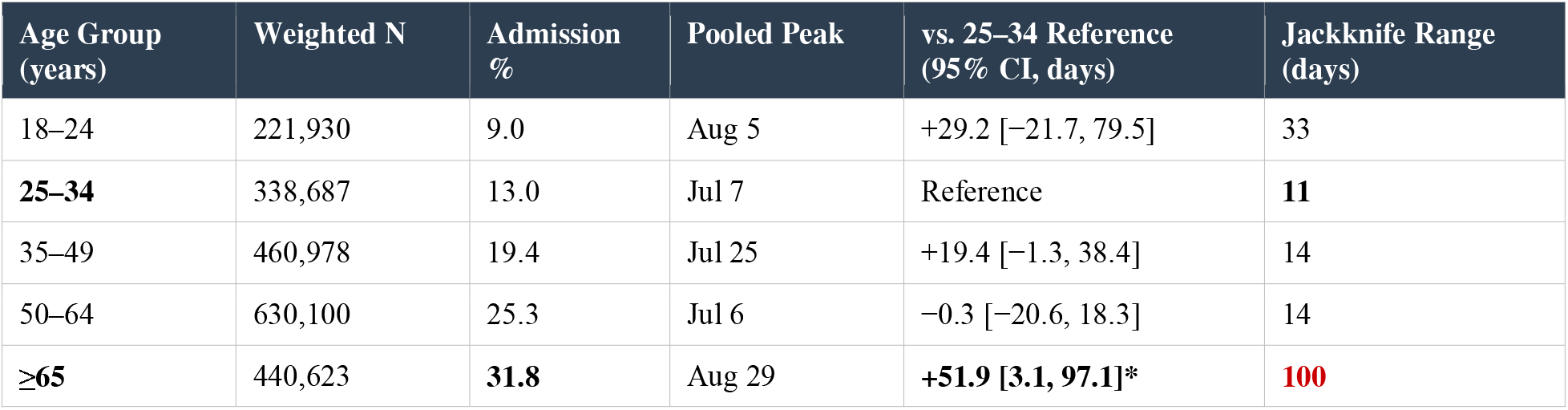
Pooled cosinor peak estimates, pairwise bootstrap comparison to reference group (25–34 years), and jackknife reproducibility range by age group. *95% CI excludes zero. Admission % = proportion resulting in hospital admission. Jackknife range for ≥65 is ∼10× larger than working-age groups.

| Age Group (years) | Weighted N | Admission % | Pooled Peak | vs. 25–34 Reference (95% CI, days) | Jackknife Range (days) |
| --- | --- | --- | --- | --- | --- |
| 18–24 | 221,930 | 9.0 | Aug 5 | +29.2 [–21.7, 79.5] | 33 |
| **25–34** | 338,687 | 13.0 | Jul 7 | Reference | **11** |
| 35–49 | 460,978 | 19.4 | Jul 25 | +19.4 [–1.3, 38.4] | 14 |
| 50–64 | 630,100 | 25.3 | Jul 6 | –0.3 [–20.6, 18.3] | 14 |
| **$\geq 65$** | 440,623 | **31.8** | Aug 29 | **+51.9 [3.1, 97.1]\*** | **100** |

### Pooled Seasonal Peak Timing by Age

The pooled likelihood ratio test confirmed a significant age × seasonal-phase interaction (χ^2^[8] = 2356.2, p < .0001). Pairwise bootstrap comparisons against the 25–34-year reference group found that only the ≥65-year group differed significantly in peak timing (+51.9 days, 95% CI: 3.1–97.1). Adults aged 18–24 (+29.2 days, 95% CI: −21.7 to 79.5), 35–49 (+19.4 days, 95% CI: −1.3 to 38.4), and 50–64 (−0.3 days, 95% CI: −20.6 to 18.3) did not differ significantly. Jackknife resampling confirmed that ages 25–64 showed stable, recurring mid-summer peaks (jackknife range: 11–14 days), while the ≥65-year group showed a 100-day jackknife range, indicating instability of the pooled estimate (Table 1).

### Sex-Stratified Results

Sex-stratified cosinor analyses revealed a striking divergence in the ≥65-year group that was not apparent in the overall analysis (Table 2; Figure 1, Panels C–E).

**Table 2.** Sex-stratified cosinor results by age group. Admission % = hospital admission rate. The 332-day jackknife range for females ≥65 years, compared with 31 days for males ≥65, indicates that seasonal instability in the oldest age group is driven by the female subgroup.

| Age Group | Weighted N | Admission % | Pooled Peak | Amplitude | Jackknife Range |
| --- | --- | --- | --- | --- | --- |
| Male |  |  |  |  |  |
| 18–24 | 135,583 | 10.9 | Jul 25 | 0.080 | $\sim 33$ days |
| 25–34 | 229,044 | 14.5 | Jul 8 | 0.197 | 11 days |
| 35–49 | 323,794 | 21.0 | Jul 25 | 0.137 | 14 days |
| 50–64 | 437,597 | 26.2 | Jul 8 | 0.132 | 14 days |
| **$\geq 65$** | 278,530 | 31.6 | Aug 9 | 0.079 | 31 days |
| Female |  |  |  |  |  |
| 18–24 | 86,201 | 6.0 | Sep 12 | 0.038 | ~wide |
| 25–34 | 109,575 | 9.7 | Jun 28 | 0.074 | stable |
| 35–49 | 136,944 | 15.5 | Jul 21 | 0.063 | stable |
| 50–64 | 192,250 | 23.2 | Jun 30 | 0.082 | stable |
| ≥65 | 162,066 | 32.1 | **Nov 28** | 0.045 | **332 days** |

**Figure 1.**
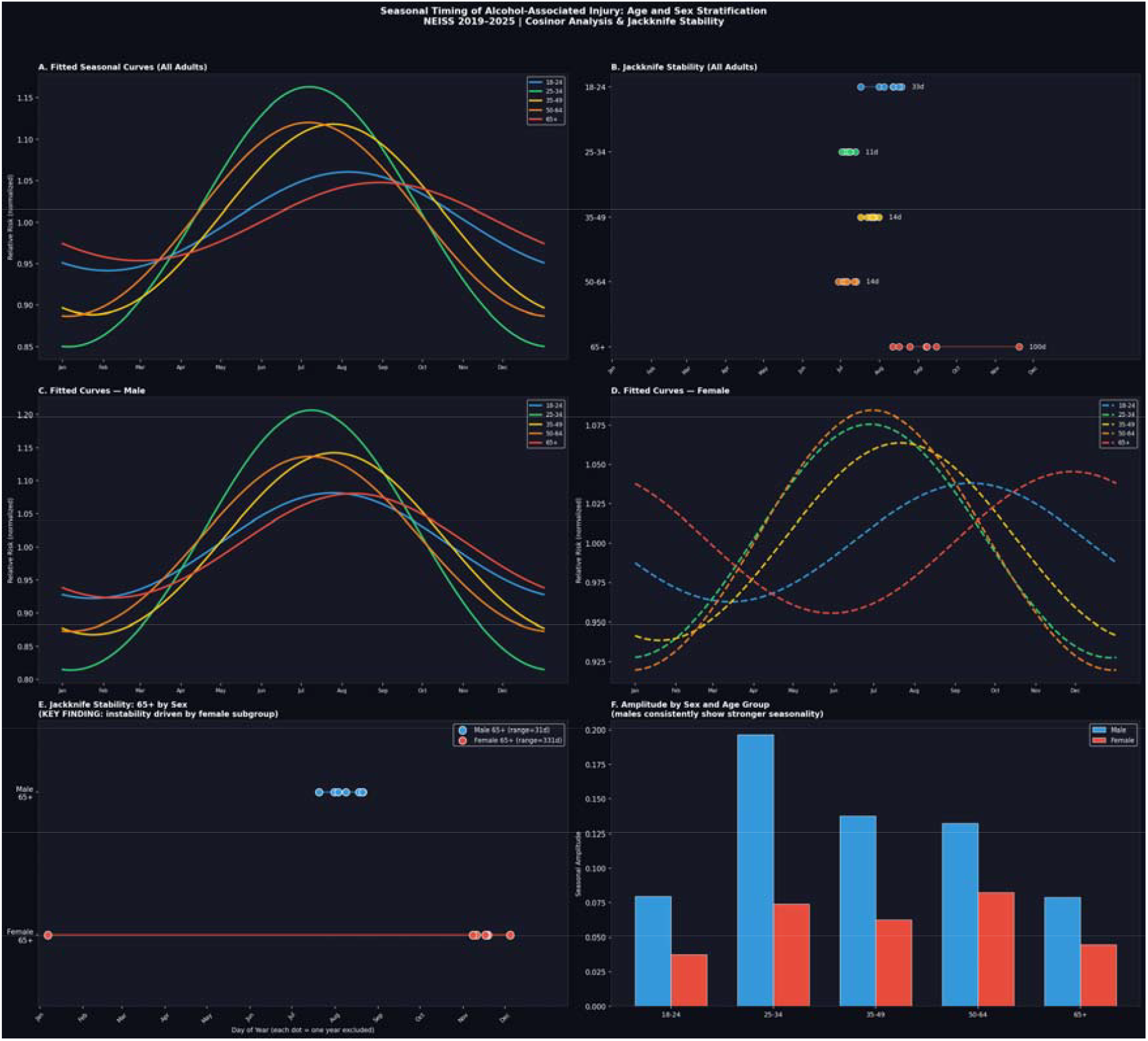
(A) Fitted cosinor curves by age group, pooled 2019–2025, normalized to each group’s annual mean. (B) Overall jackknife stability by age group. (C) Male-stratified fitted curves. (D) Female-stratified fitted curves. (E) Jackknife stability comparison for the ≥65 group by sex: males show a tight August cluster (31-day range); females show near-complete calendar-year instability (332-day range). (F) Seasonal amplitude by sex and age group.

Among males, seasonal peaks across all age groups fell within a tight July–August window (July 8 to August 9), with jackknife ranges ≤33 days in every group including ≥65 years (jackknife range: 31 days, peak consistently in early August across all seven jackknife iterations). Males aged ≥65 thus show a reproducible seasonal pattern essentially indistinguishable in stability from working-age males.

Among females, working-age groups (25–64 years) also showed stable mid-summer peaks (June 28 to July 21) comparable to their male counterparts, with jackknife ranges of similar magnitude. However, females aged ≥65 years showed a pooled peak in late November and a jackknife range spanning 332 days — effectively the entire calendar year. Individual-year jackknife estimates for this stratum ranged from early November through early January, with no single year’s exclusion shifting the estimate into summer. Importantly, seasonal variation was statistically significant in every individual year for this group (all p < .0001), and seasonal amplitude (mean = 0.045) was lower than in all other age-sex strata, suggesting that older women’s seasonal injury pattern is real but diffuse, low-amplitude, and concentrated in autumn through winter rather than summer.

## DISCUSSION

In this analysis of seven years of nationally representative U.S. ED surveillance data, we found that adults aged 25–64 years exhibit a reproducible, well-localized mid-summer peak in alcohol-associated injury that recurs reliably from year to year. The previously reported temporal instability among adults ≥65 years overall was shown, upon sex stratification, to be driven entirely by the female subgroup: older males showed a highly stable August peak (jackknife range: 31 days), while older females showed peak timing that was effectively undetermined, spanning nearly the entire calendar year across jackknife iterations (332 days), with estimates consistently concentrated in autumn and early winter. To our knowledge, this is the first study to identify this sex-specific pattern in alcohol-associated injury seasonality, and the first to apply leave-one-year-out jackknife resampling to this clinical surveillance question.

This finding has direct clinical and public health relevance. First, hospital admission rates in our sample increased more than three-fold across the age range studied (9.0% in adults 18–24 years to 31.8% in adults ≥65 years), consistent with the well-described amplification of alcohol injury severity by age-related physiologic vulnerability, polypharmacy, and comorbid disease (Centers for Disease Control and Prevention, 2022; Saitz, 2005). The combination of the most severe injury outcomes and the least predictable seasonal risk window in older women specifically marks this subgroup as particularly difficult to address with conventional, calendar-based prevention approaches.

Several mechanisms may explain the sex-specific pattern in older adults. Older women’s alcohol-related injury risk may be more strongly influenced by factors that vary year to year without a fixed calendar rhythm — including changes in social isolation, medication regimens, caregiving roles, and age-related physiological changes in alcohol sensitivity that interact with seasonal temperature, lighting, and activity patterns (Nolen-Hoeksema, 2004). It is also possible that the autumn and early winter peak in older women reflects a distinctive pattern of holiday-related or cold-weather-associated alcohol use among this population, perhaps linked to social gatherings, seasonal depression, or reduced outdoor activity, that differs fundamentally from the summer-outdoor-activity pattern driving peaks in younger adults and older men (Saitz, 2005; Merikangas et al., 1990).

The practical implication for ED practitioners is that fixed-calendar alcohol screening intensification ( e.g., heightened AUDIT-C screening or alcohol-use counseling during summer months) is appropriate for working-age adults of both sexes and for older men. For older women presenting to the ED with fall-related or other traumatic injury, year-round structured alcohol screening (regardless of season) appears warranted, as no reliable calendar window can be identified for concentration of prevention efforts in this group.

### Limitations

NEISS captures only ED-treated injuries, excluding outpatient, EMS-only, and prehospital deaths. Alcohol involvement depends on clinical documentation and testing, which may vary across hospitals, over time, and by patient age and sex (if clinicians test less consistently for alcohol in older women presenting with falls, the already-lower case count in this stratum would further reduce power and widen the jackknife range). Pooling all injury mechanisms may mask mechanism-specific seasonal patterns, particularly falls, which predominate in the ≥65-year group. Seven years provides limited replicates for jackknife analysis. This ecological design cannot establish individual-level mechanisms.

### Future Directions

Stratified analyses by injury mechanism (falls versus motor vehicle crashes versus assaults) would clarify whether the sex-age interaction in peak timing is driven by specific injury types. Extending the surveillance window, linking to regional climate and social calendar data, and integrating age- and sex-specific alcohol consumption data from national health surveys (e.g., NHANES) could test specific behavioral mechanisms. Future studies recruiting older female patients from ED settings to assess alcohol use patterns and seasonality prospectively would directly address the most clinically urgent subgroup identified here.

## CONCLUSIONS

Using seven years of nationally representative U.S. ED surveillance data and a novel jackknife reproducibility framework, we found that alcohol-associated injury seasonality is reproducible and summer-concentrated in working-age adults of both sexes and in older men, but is temporally non-stationary and diffuse in older women, who also experience disproportionately severe injury outcomes. These findings argue for fixed-calendar prevention targeting July–August in working-age adults and older men, and for year-round vigilance in older women, for whom no reliable seasonal risk window can be identified at the population level.

## Data Availability

All source data were openly available before the initiation of the study and were accessible at cpsc.gov/Research--Statistics/NEISS-Injury-Data. All data produced in the present study are available upon reasonable request to the authors.

## ACKNOWLEDGEMENTS

We would like to thank Dr. Cheryl Cherpitel for her highly valued mentorship in this project.

